## Supplementary figures and tables for "Delayed generation of functional virus-specific circulating T follicular helper cells correlates with severe COVID-19"

Supplementary figure 1

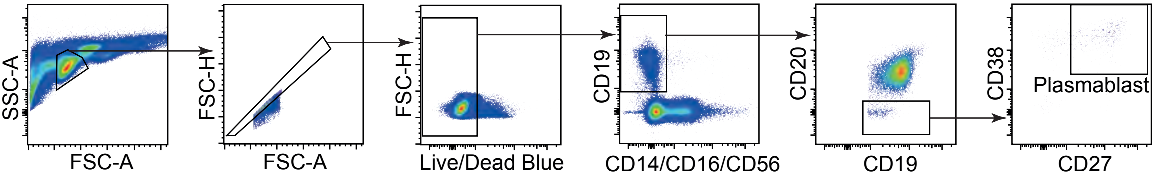


**Representative plasmablast gating strategy.** Representative gating strategy to identify plasmablast in PBMC by flow cytometry. B cells were identified with lineage (CD14/CD16/CD56) negative but CD19+ population. Within CD20 negative population, cells expressing CD27 and CD38 were identified as plasmablast.

Supplementary figure 2

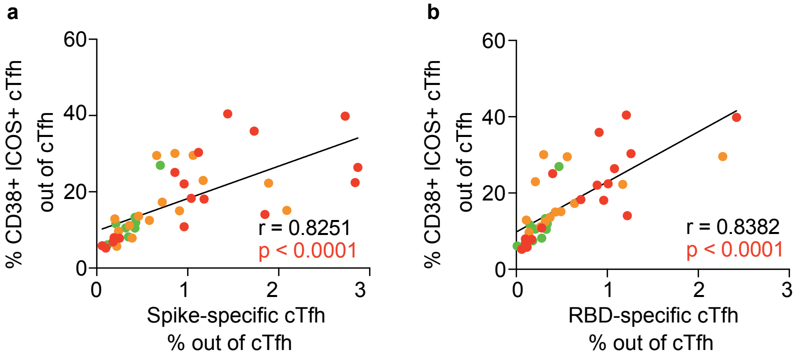


**Frequency of activated cTfh cells correlated with frequency of virus-specific cTfh cells during acute disease.** Spearman correlation for frequency of (a) spike-specific cTfh cells and (b) RBD-specific cTfh cells versus frequency of CD38+ ICOS+ cTfh cells. The dots are color-coded according to peak disease severity. 9, 14 and 18 individual samples from 9 mild, 14 moderate and 18 severe patients were analysed. For patients with longitudinal acute samples, data from the earliest sample was involved as representative in Spearman correlation analysis.

Supplementary figure 3

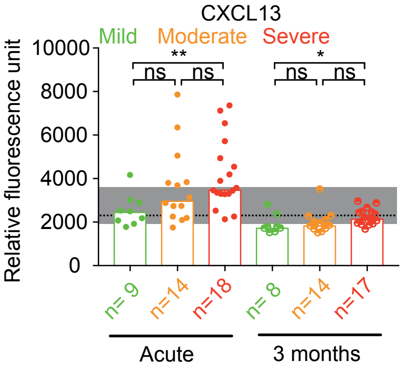


**Level of plasma CXCL13 in COVID-19 patients during acute disease and 3 months convalescence.** Bar charts show the relative fluorescence unit (RFU) of plasma CXCL13 with median from COVID-19 patients with acute infection (Acute) and 3 months convalescence (3 months). Dotted line shows the median frequency with 95% CI (grey area) of healthy controls. For patients with longitudinal acute samples, data from the earliest sample was involved as representative in Spearman correlation analysis. Kruskal-Wallis with Dunn's multiple comparisons test was used to assess statistical significance at p<0.05. ** p<0.01.

Supplementary figure 4

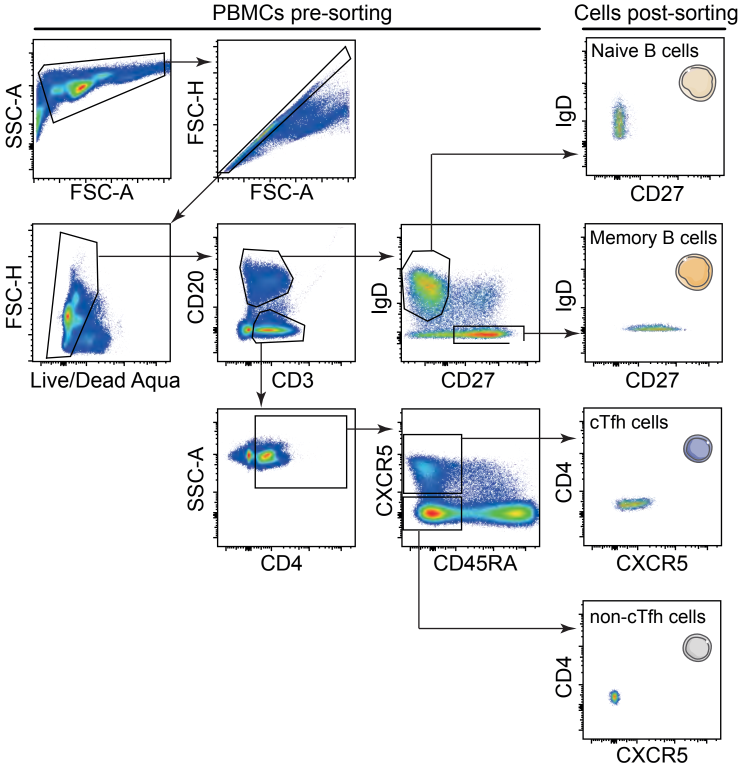


**Representative sorting strategy of cTfh, non-cTfh, memory and naïve B cells.** Representative sorting strategy to isolate cTfh, non-cTfh, memory and naïve B cells from PBMC by flow cytometry. From single, live CD20- but CD3+ CD4+ T cells, memory CD4+ T cells were identified as CD45RA-. From memory CD4+ T cells, CXCR5+ cells were sorted and identified as cTfh cells and CXCR5- cells were sorted and identified as non-cTfh cells. From single, live CD3- but CD20+ B cells, CD27- but IgD+ and CD27+ but IgD- cells were sorted and identified as naïve and memory B cells respectively.

Supplementary figure 5

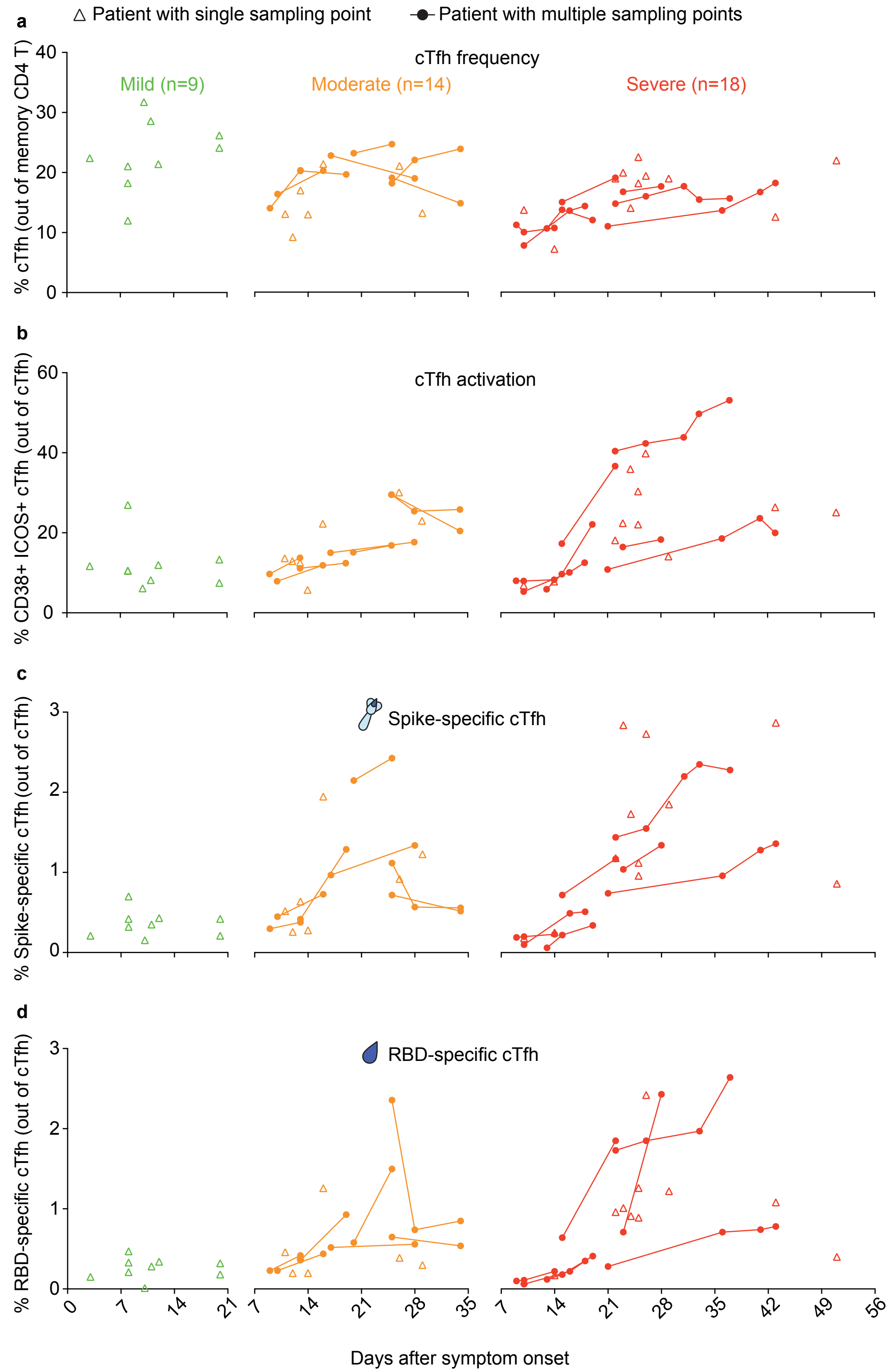

Supplementary figure 5

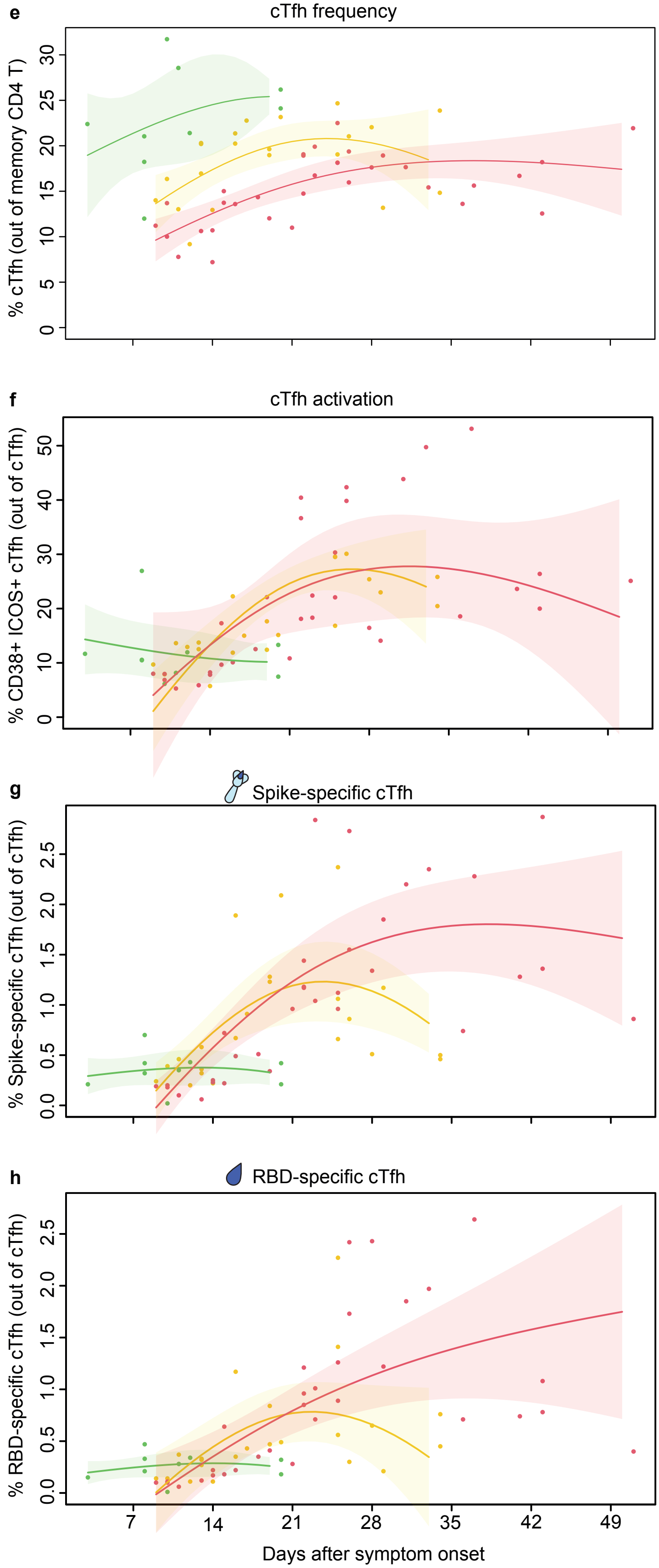


**Longitudinal distribution of cTfh cells in individual COVID-19 patients across disease severity during acute infection.** (a-d) Figures show the longitudinal distribution of frequencies of (a) cTfh cells (b) activated cTfh cells (c) Spike-specific cTfh cells and (d) RBD-specific cTfh cells in individual COVID-19 patients across disease severity during acute infection. The dots are color-coded according to peak disease severity. Open triangles show patients with single sampling point, and dots linked with line show patients with multiple, longitudinal sampling points. (e-h) Graphs show frequencies of (e) cTfh cells (f) activated cTfh cells (g) Spike-specific cTfh cells and (h) RBD-specific cTfh cells COVID-19 patients over time. The dots and lines are color-coded according to peak disease severity. Lines show group mean estimates based on one GEE model using restricted cubic splines for the time effect and group time interaction for the estimation of group differences. The shaded areas are 95% CI.

Supplementary figure 6

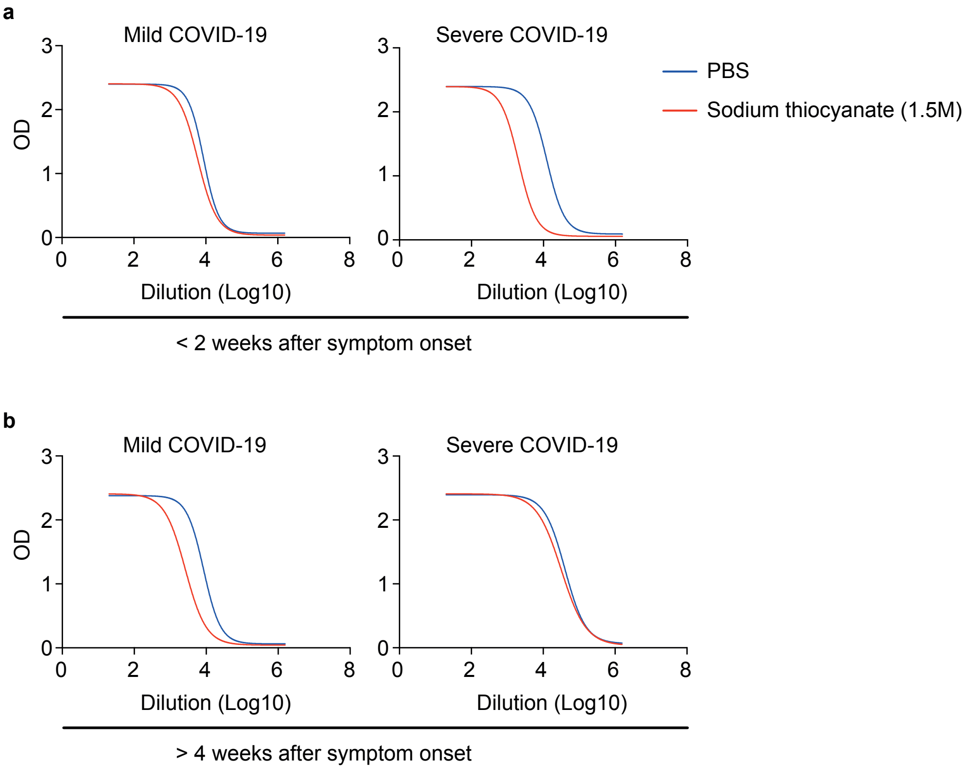


**Representative measurement of plasma IgG against SARS-CoV-2 spike avidity by ELISA.** Representative plasma samples from mild and severe COVID-19 patients with symptom onset (a) less than 2 weeks and (b) over 4 weeks were tested by ELISA to determine the avidity of IgG against SARS-CoV-2 spike. Representative binding curves for plasma IgG against spike by ELISA in a 5-fold dilution series starting from 1:20 to 1:312500 and incubating with (Blue curve) PBS or (Red curve) 1.5M solution of sodium thiocyanate. EC50 have been calculated using non-linear fit of the data to a sigmoidal curve by constraining the top of each curve to the OD of 2.5.

Supplementary figure 7

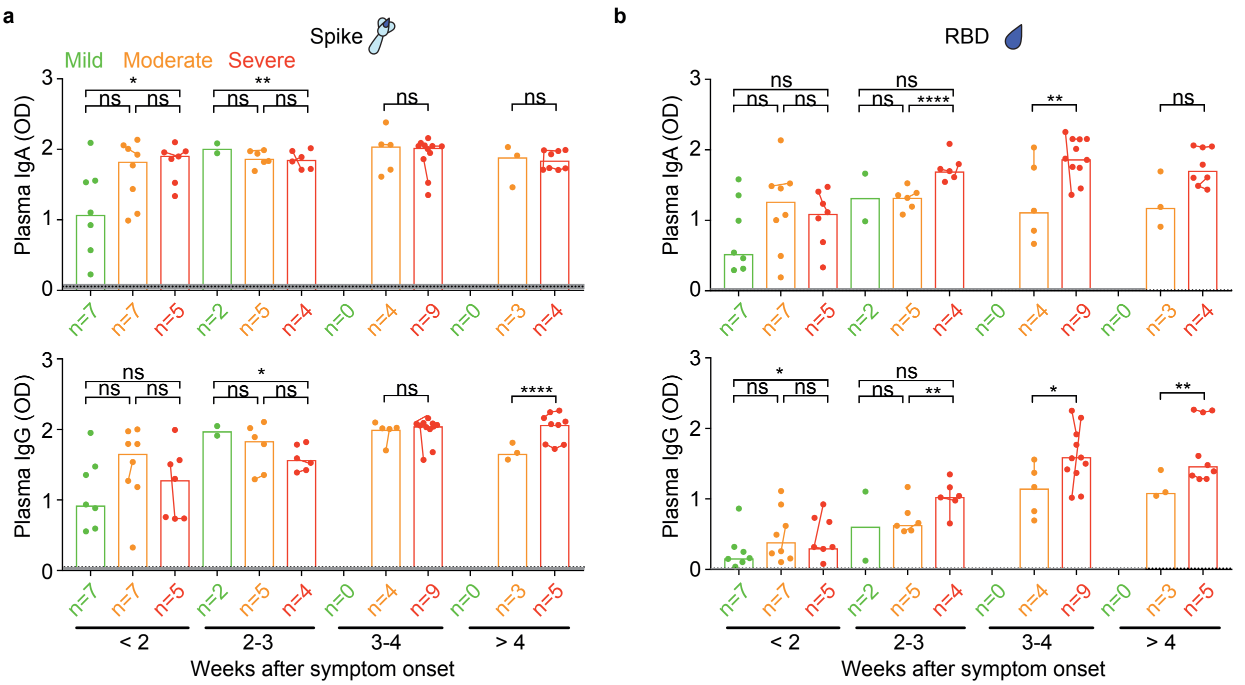


**Titers of plasma immunoglobulins against SARS-CoV-2 spike and RBD according to weeks after symptom onset.** Titers of plasma IgA and IgG against SARS-CoV-2 (a) spike and (b) RBD were tested by ELISA and bar charts show the OD value with median in COVID-19 patients with acute disease according to weeks after symptom onset. Dots are individual samples color-coded according to peak disease severity. 9, 22 and 33 individual samples from 9 mild, 14 moderate and 18 severe patients respectively were analysed. X axis shows number of patients in each bar. One severe patient displaying IgA deficiency was excluded in all IgA analysis. Dots from same patient are linked with line in each bar. Dotted lines show the median frequency with 95% CI (grey area) of healthy controls. Time-period specific differences between groups were calculated using by using time-period specific subsets of the data. The graphical presentations of the different outcomes were based on a GEE model with time modelled using a restricted cubic spline with knots at 0, 14, 21, 28 and 53 days. Statistically significant differences were assessed at p<0.05, ** p<0.01, *** p<0.001, **** p<0.0001.

Supplementary figure 8

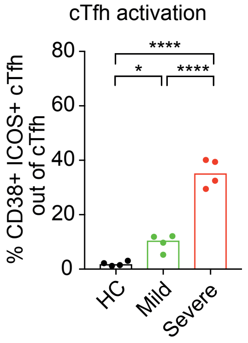


**Frequency of activated cTfh cells in COVID-19 patients and healthy donors selected in cTfh functional assay.** Bar charts show the frequency of activated cTfh cells with mean in COVID-19 patients with acute disease and healthy donors selected in cTfh functional assay. One-Way ANOVA was used to consider statistically significant at $p<0.05$. ** $p<0.01$, *** $p<0.001$, **** $p<0.0001$.

Supplementary figure 9

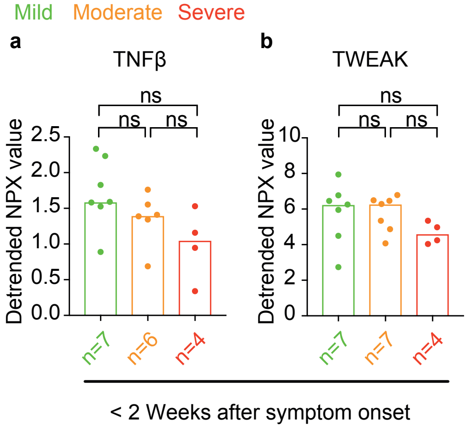


**Levels of plasma TNFβ and TWEAK in COVID-19 patients sampled during early SARS-CoV-2 infection (less than 2 weeks after symptom onset).** Bar charts show the detrended NPX value of plasma (a) TNFβ and (b) TWEAK with median from COVID-19 patients during less than 2 weeks after symptom onset. For patients with longitudinal acute samples, data from the earliest sample was involved as representative. X axis shows number of patients in each bar. Kruskal-Wallis with Dunn's multiple comparisons test was used to assess statistical significance at $p<0.05$.

Supplementary Table 1. Flow cytometry panel of cTfh cells phenotyping

| Phenotyping of cTfh cells | | | |
|---|---|---|---|
| Fluorochrome | Marker | Company | Clone |
| PerCP-Cy5.5 | CD4 | Biolegend | OKT4 |
| FITC | CD62L | Biolegend | DREF-56 |
| PE-Cy7 | ICOS | Biolegend | C398-4A |
| PE-Cy5 | CD40L | Biolegend | 24-31 |
| PE-TR | CXCR5 | Invitrogen | MU5UBEE |
| PE | CD38 | Biolegend | HIT2 |
| APC-Cy7 | CD3 | BD Biosciences | SK7 |
| AF700 | CCR7 | Biolegend | G043H7 |
| BV786 | CCR6 | BD Biosciences | 11A9 |
| BV650 | CXCR3 | Biolegend | G025H7 |
| BV421 | PD1 | BD Biosciences | EH12.1 |
| DAPI | L/D Blue | Invitrogen | cat no. L34962 |
| BUV395 | CD45RA | BD Biosciences | HI100 |

Supplementary Table 2. Flow cytometry panel of B cells phenotyping

| Phenotyping of B cells | | | |
|---|---|---|---|
| Fluorochrome | Marker | Company | Clone |
| PerCP-Cy5.5 | CD38 | BD Biosciences | HIT2 |
| PE-Cy7 | CD19 | BD Biosciences | HIB19 |
| APC-Cy7 | CD20 | BD Biosciences | 2H7 |
| BV786 | IgG | BD Biosciences | G18-145 |
| BV650 | CD27 | BD Biosciences | M-T271 |
| BV510 | CD14 | Biolegend | M5E2 |
| BV510 | CD16 | BD Biosciences | 3G8 |
| BV510 | CD56 | BD Biosciences | B159 |
| Indo I Violet | L/D blue | Invitrogen | cat no. L34962 |
| BUV 395 | IgM | BD Biosciences | G20-217 |
| BUV 395 | IgD | BD Biosciences | IA6-2 |

Supplementary Table 3. Flow cytometry panel of SARS-CoV-2-specific cTfh cells phenotyping

| Phenotyping of SARS-CoV-2-specific cTfh cells | | | |
|---|---|---|---|
| Fluorochrome | Marker | Company | Clone |
| PerCP-Cy5.5 | CD134 | BD Biosciences | ACT35 |
| PE-TR | CXCR5 | Invitrogen | MU5UBEE |
| APC-Cy7 | CD3 | BD Biosciences | SK7 |
| AF700 | CD4 | BD Biosciences | L200 |
| APC | CD25 | Biolegend | BC96 |
| BV786 | CCR6 | BD Biosciences | 11A9 |
| BV650 | CXCR3 | Biolegend | G025H7 |
| DAPI | L/D Blue | Invitrogen | cat no. L34962 |
| BUV395 | CD45RA | BD Biosciences | HI100 |

Supplementary Table 4. Flow cytometry panel of cTfh and B cells sorting

| cTfh and B cells sorting | | | |
|---|---|---|---|
| Fluorochrome | Marker | Company | Clone |
| FITC | IgD | BD Biosciences | IA6-2 |
| PE | CD3 | BD Biosciences | SK7 |
| PE-Cy5 | CD45RA | BD Biosciences | 5H9 |
| PE-TR | CXCR5 | Invitrogen | MU5UBEE |
| APC-Cy7 | CD20 | BD Biosciences | 2H7 |
| AF700 | CD4 | BD Biosciences | L200 |
| BV650 | CD27 | BD Biosciences | M-T271 |
| BV510 | L/D Aqua | Invitrogen | cat no. L34966 |

Supplementary Table 5. Demographic and clinical information of COVID-19 patients involved in *in vitro* co-culture assay.

| Peak Disease Severity | Mild | Severe | Significance [A] |
|---|---|---|---|
| Number of Individuals | 4 | 4 | |
| Age in years, mean ± SD | 55 ± 3.7 (51-60) | 68 ± 5.4 (24-81) | <0.01 |
| Male, n (%) | 2 (50) | 4 (100) | ns |
| Female, n (%) | 2 (50) | 0 (0) | |
| Days after onset of stmptom, mean± SD (range) | 11 ± 1.3 (9-12) | 34 ± 14.6 (18-53) | <0.05 |

[A] Mann-Whitney U unpaired t-test was performed to determine statistical significance.
